# Agreement between commercially available ELISA and in-house Luminex SARS-CoV-2 antibody immunoassays

**DOI:** 10.1101/2021.03.09.21252401

**Authors:** Rebeca Santano, Diana Barrios, Fàtima Crispi, Francesca Crovetto, Marta Vidal, Jordi Chi, Luis Izquierdo, Eduard Gratacós, Gemma Moncunill, Carlota Dobaño

## Abstract

Serological diagnostic of the severe respiratory distress syndrome coronavirus 2 (SARS-CoV-2) is a valuable tool for the determination of immunity and surveillance of exposure to the virus. In the context of an ongoing pandemic, it is essential to externally validate widely used tests to assure correct diagnostics and epidemiological estimations. We evaluated the performance of the COVID-19 ELISA IgG and IgM/A (Vircell, S.L.) against a highly specific and sensitive in-house Luminex immunoassay in a set of samples from pregnant women and cord blood. The agreement between both assays was moderate to high for IgG but low for IgM/A. Considering seropositivity by either IgG and/or IgM/A, the technical performance of the ELISA was highly imbalanced, with 96% sensitivity at the expense of 22% specificity. As for the clinical performance, the negative predictive value reached 87% while the positive predictive value was 51%. Our results stress the need for highly specific and sensitive assays and external validation of diagnostic tests with different sets of samples to avoid the clinical, epidemiological and personal disturbances derived from serological misdiagnosis.

## Introduction

In the context of an ongoing pandemic, the importance of accurate diagnostic methods for disease control and elimination has been underpinned. Coronavirus disease 2019 (COVID-19) serological diagnostic based on antibody detection against the severe respiratory distress syndrome coronavirus 2 (SARS-CoV-2) is a valuable tool that allows for the determination of immunity development and surveillance of exposure to the virus (1,2). In the case of COVID-19, it is thought that antibodies mediate protection via a myriad of functions. (3). Therefore, good serological tests are required not only for epidemiological surveillance and policy implementation, but also for helping elucidate the mechanisms involved in protection and the susceptibility to reinfection after exposure to the virus (1,2). Additionally, serological tests may be also useful for establishing vaccine induced protection and finding blood donors that qualify to obtain plasma to be used as treatment for severe COVID-19 patients (1,2,4).

The definition of a good serological test in terms of technical performance is based on the values of specificity (SP) and sensitivity (SE). The clinical relevance of a serological test is defined by the negative (NPV) and positive (PPV) predictive values. These last two parameters depend on the prevalence of the disease, while theoretically SP and SE do not. Since achieving a high score of both SP and SE is generally difficult, prioritizing one of them during the test development process has personal, social and clinical implications, especially in the context of a pandemic where prevalence of disease might not be correctly defined.

Several antibody detection tests for SARS-CoV-2 are available in the market, such as rapid diagnostic tests (RDT), enzyme-linked immunosorbent assays (ELISA), neutralization assays, and chemiluminescent immunoassays (CLIA) (5). Here we compared the performance of a commercially available COVID-19 ELISA (Vircell Microbiologists, Granada, Spain) with the performance of an in-house fluorescence-based, high-throughput and multiplex Luminex immunoassay (6). The Vircell COVID-19 ELISA (from now on ELISA) detects specific IgG or IgM and IgA together (IgM/A) against the nucleocapsid (N) and spike (S) antigens adsorbed on solid phase (7). The Luminex immunoassay has been optimized for the detection of specific IgG, IgM and IgA separately against a multiplex panel of 5 antigens adsorbed on magnetic beads that are in suspension (6).

Thanks to the simplicity of the method and its automatization, ELISA is used in the clinical practice (8–11). Internal validation from Vircell, S.L. reports a 88% SE and 99% SP for IgM/A assay and a 85% SE and 98% SP for the IgG assay (7). External validation of widely used tests during this COVID-19 pandemic is essential to assure correct diagnostics and epidemiological estimations. We have previously reported that the in-house Luminex assay reaches up to 100% SP and 95.78% SE (6). Our aim is to report the performance of the ELISA IgG and IgM/A externally assessed using our highly specific and sensitive Luminex immunoassay.

## Methods

### Study population and sample selection

We analyzed 283 samples from peripheral blood from pregnant women and cord blood (**Table 1**). Of those, 168 mothers belonged to a larger cohort of pregnant women whose samples were collected in the context of a study focused on the effects of COVID-19 on pregnancy outcomes. The study population and sample collection methods have been described elsewhere (12). The selection of samples in this report was based on the results obtained by ELISA and the suspicion of low SP by the IgM/A assay. In particular, it was the detection of 3 IgG and IgM/A positive tests in cord blood samples paired with mothers who had evidence of past and/or present infection that led to further investigation. While IgG crosses the placenta efficiently, a very limited amount of IgA passes from mother to fetus and IgM does not cross the placenta during normal pregnancies (13). As a result, the presence of an IgM/A positive test suggested congenital COVID-19 or false-positive results. Therefore, the selection included the aforementioned IgG+ and IgM/A+ cord blood samples (n= 3) together with IgG-and IgM/A-(n= 23), IgG+ and IgM/A+ (n= 22), and IgG- and IgM/A+ (n= 123) samples from pregnant women. The remaining samples were from healthy pre-pandemic pregnant women (n= 73) and cord blood (n= 39) (**Table 1**). Samples analyzed in this study received ethical clearance for immunological evaluation by the Review Board at each institution.

**Table 1.**
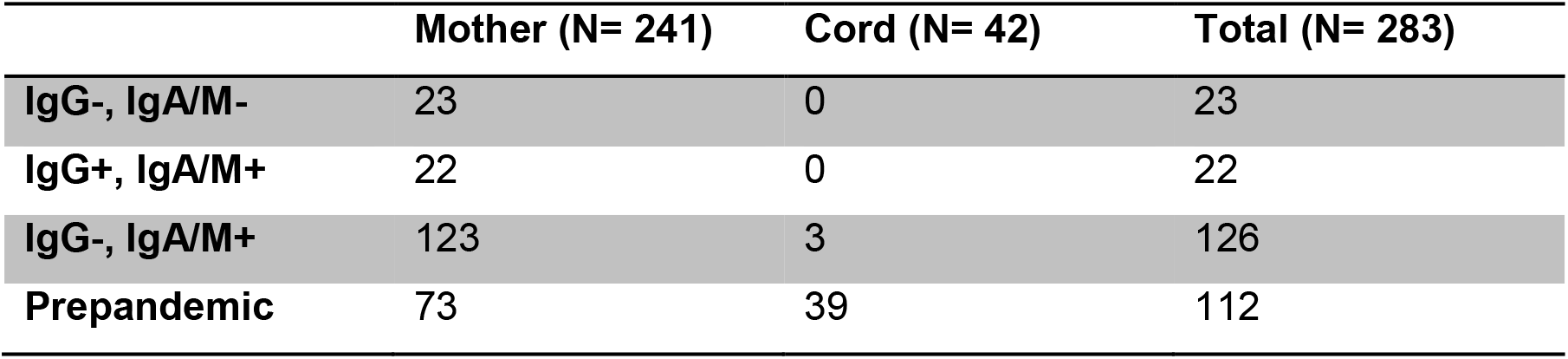
Sample size and classification of samples according to their biological origin and ELISA results.

|  | Mother (N= 241) | Cord (N= 42) | Total (N= 283) |
| --- | --- | --- | --- |
| **IgG-, IgA/M-** | 23 | 0 | 23 |
| **IgG+, IgA/M+** | 22 | 0 | 22 |
| **IgG-, IgA/M+** | 123 | 3 | 126 |
| **Prepandemic** | 73 | 39 | 112 |

### ELISA IgG and IgM/A

The IgG and IgM/A assays were performed as indicated by the manufacturer. Briefly, samples were diluted and, in the case of the IgM/A assay, they were then incubated with an IgG sorbent to eliminate IgG from plasma and any possible interference. Then, in both assays, samples and controls from the kit were incubated at 37°C for 45 min. After a washing step, they were incubated with peroxidase-conjugated detection antibodies (anti-human IgG or anti-human IgM+IgA) at 37°C for 30 min. After another wash, they were incubated with substrate solution for 20 min in the dark. Finally, stop solution was added and the optical density was read at 450 nm. The cut-off value was established based on the manufacturer’s procedure. The ELISA IgG and IgM/A assays are based on the detection of specific antibodies against the N and S antigens adsorbed on a solid surface. Quantification is based on optical density after the reaction of the enzyme-linked secondary antibodies in contact with a substrate, and it is detected in a spectrophotometer.

### Luminex

The in-house Luminex immunoassay has been optimized for the detection of specific IgG, IgM and IgA separately against a multiplex panel of antigens adsorbed on magnetic beads that are in suspension (6). Each magnetic bead region is characterized by a unique mix of two fluorochromes that allows their identification by laser excitation. Each antigen is coupled to a specific bead region and the multiplex panel included: the full-length N (N FL) antigen, a specific C-terminal region of N (N CT) (14), the full-length S, the subunit 2 from the S antigen (S2) and the receptor binding domain (RBD) in S1. Quantification is based on the detection of fluorescence emission by a phycoerythrin-labelled secondary antibody and it is detected in a Luminex FLEXMAP 3D instrument.

## Results and Discussion

### Moderate to high agreement between specific IgG measured by ELISA and Luminex

Most of the IgG positive samples detected by ELISA were also classified as IgG positive by each of the antigens included in the Luminex panel (**Figure 1 and Table 2**), and many of the negative samples classified by ELISA were also classified as negative by each of the antigens included in the Luminex panel.

**Table 2.** Classification of samples by ELISA and Luminex immunoassays for specific IgG, IgM and IgA and percentage of agreement between the tests. The table shows the number of samples classified by the two tests taking into account specific IgG, IgM and IgA against any of the antigens included in the Luminex multiplex panel or the combination of all of them. Concordant samples are those classified as negative (LM-E-) or positive (LM+ E+) by both tests, while discrepant samples are those with opposite results in both tests (LM+ E- and LM-E+). The percentages of agreement were calculated by dividing the concordant positive samples (LM+ E+) or concordant negative samples (LM-E-) by the total of samples that tested positive (LM+ E+, LM+ E-, LM-E+) or negative (LM-E-, LM+ E-, LM-E+) in at least one of the tests, respectively. LM: Luminex, E: ELISA.

| Isotype | Antigen | LM -<br>E - | LM -<br>E + | LM +<br>E - | LM +<br>E + | Positive<br>agreement | Negative<br>agreement |
| --- | --- | --- | --- | --- | --- | --- | --- |
| IgG | N FL | 142 | 10 | 1 | 18 | 62.07% | 92.81% |
|  | N CT | 130 | 5 | 13 | 23 | 56.10% | 87.84% |
|  | S | 132 | 1 | 11 | 27 | 69% | 91.67% |
|  | S2 | 142 | 1 | 1 | 27 | 93.10% | 98.61% |
|  | RBD | 133 | 1 | 10 | 27 | 71.05% | 92.36% |
|  | Combination | 121 | 1 | 22 | 27 | 54.00% | 84.03% |
| IgM | N FL | 25 | 122 | 0 | 24 | 16.44% | 17.01% |
|  | N CT | 25 | 139 | 0 | 7 | 4.79% | 15.24% |
|  | S | 25 | 127 | 0 | 19 | 13.01% | 16.45% |
|  | S2 | 24 | 134 | 1 | 12 | 8.22% | 15.19% |
|  | RBD | 25 | 132 | 0 | 14 | 9.59% | 15.92% |
|  | Combination | 24 | 112 | 1 | 34 | 23.29% | 17.65% |
| IgA | N FL | 25 | 121 | 0 | 25 | 17.12% | 17.12% |
|  | N CT | 25 | 133 | 0 | 13 | 8.90% | 15.82% |
|  | S | 22 | 119 | 3 | 27 | 18.49% | 15.60% |
|  | S2 | 23 | 115 | 2 | 31 | 21.23% | 16.67% |
|  | RBD | 23 | 121 | 2 | 25 | 17.12% | 15.97% |
|  | Combination | 22 | 102 | 3 | 44 | 30.14% | 17.74% |

**Figure 1.**
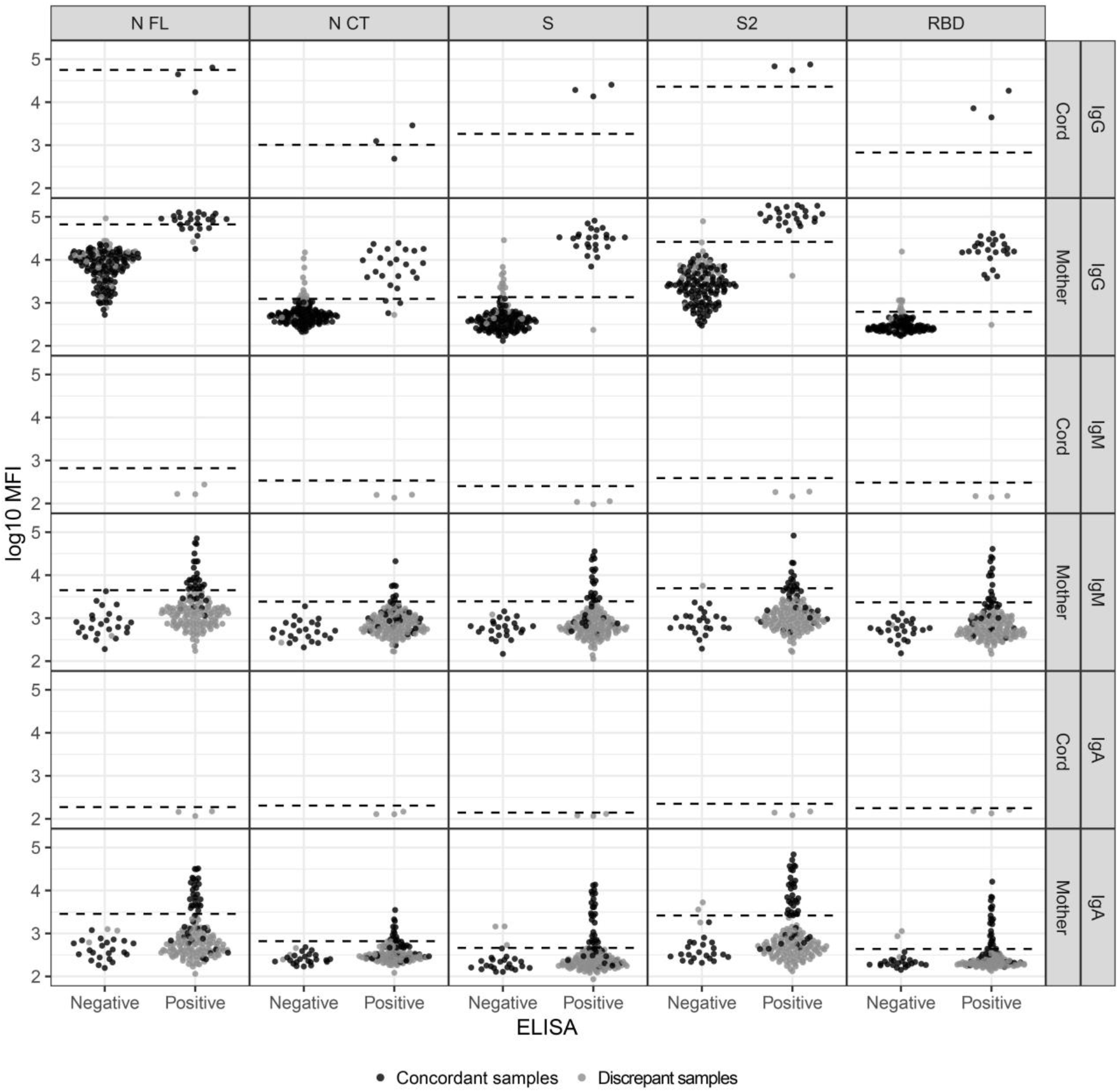
Classification of samples by ELISA and Luminex immunoassays for specific IgG, IgM and IgA. The categories in the X-axis refer to the classification of the ELISA assay (negative or positive). The Y-axis indicates the Luminex assay result in log_10_ transformed median fluorescence intensity (MFI). The seropositivity cutoff is shown in dashed lines (mean plus 3 SD of prepandemic samples). The color classification in concordant (black) vs. discordant (grey) sample results between Luminex and ELISA tests is based on the combination of all antigens within each isotype for Luminex. That is, seropositivity and seronegativity was calculated per each immunoglobulin with the combination of all antigens from the Luminex assay, and those were compared with the ELISA result to subsequently define concordant and discordant samples between tests. The figure illustrates the lower specificity of the ELISA assay for IgM/A (elevated number of false positives) compared to the Luminex IgA and IgM assays, and the higher sensitivity of the IgG Luminex for the detection of positive samples that had been classified as negative by ELISA.

For IgG, the percentage of agreement ranged from 56.10% for N CT up to 93.10% for S2 for positive samples, and from 87.84% for N CT up to 98.61% for S2 for negative samples (**Table 2**). The antigen from the Luminex panel with the highest percentage of both positive and negative agreement for IgG was S2, which is part of the S antigens included in the ELISA.

Considering the classification obtained by the combination of IgG responses against all antigens included in the Luminex multiplex panel, the positive agreement was 54% while the negative agreement was 84.03% (**Table 2**). This resulted in 23 samples (all from mothers) with discrepancy for IgG: 1 sample Luminex negative and ELISA positive (LM-E+) and 22 samples Luminex positive and ELISA negative (LM+E-) (**Figure 1** and **Table 2**). Among these discrepant samples, 19 out of the 22 LM+E-samples were asymptomatic and either PCR negative or were not PCR tested, 1 sample was asymptomatic and PCR positive, 1 sample was symptomatic 1-2 months prior to sample collection and PCR negative, and 1 sample was symptomatic during the previous 7 days to sample collection and PCR positive. The only LM-E+ sample was asymptomatic and PCR negative. The 3 cord blood samples had concordant results between Luminex and ELISA. This most certainly reflects transplacental transfer of maternal IgG, since the 3 mothers had detectable levels of specific IgG detected by both tests and IgG is able to cross the placenta (13).

Since the in-house Luminex assay has previously been assessed and it demonstrated an excellent performance (6), we used the results obtained by Luminex as the gold standard to evaluate the performance of ELISA (**Table 3)**. The usage of Luminex as gold standard is further supported by the inclusion of several antigenic regions and three immunoglobulin isotypes that allow capturing a variety of immunological responses that cannot be attained by a PCR test, for example. Therefore, we evaluated the ELISA serological test based on a serological reference (Luminex) assay. For IgG, the SE was 55%, while the SP was 99%. As for the PPV and NPP, the percentages were 96% and 85%, respectively. Taking into account only the subjects with a PCR test done (N= 110), for IgG, the SE was 62%, while the SP was 90%. As for the PPV and NPP, the percentages were 44% and 95%, respectively.

**Table 3.** Performance of ELISA based on Luminex diagnostic as gold standard. The analysis included samples from mothers (N=168) and cord (N=3). The section of “All isotypes” refers to the performance based on seropositivity by any of the three isotypes. FN: False negative; FP: False positive, TN: True negative; TP: True positive; SE: Sensitivity; SP: Specificity, PPV: Positive predictive value; NPV: Negative predictive value.

| Isotype | FN | FP | TN | TP | Total | SE (%) | SP (%) | PPV(%) | NPV(%) |
| --- | --- | --- | --- | --- | --- | --- | --- | --- | --- |
| **IgG** | 22 | 1 | 121 | 27 | 171 | 55 | 99 | 96 | 85 |
| **IgM** | 1 | 112 | 24 | 34 | 171 | 97 | 18 | 23 | 96 |
| **IgA** | 3 | 102 | 22 | 44 | 171 | 94 | 18 | 30 | 88 |
| **All isotypes** | 3 | 73 | 20 | 75 | 171 | 96 | 22 | 51 | 87 |

### Low agreement between specific IgA/M measured by ELISA and IgA and IgM measured by Luminex

Almost all the IgM/A negative samples detected by ELISA were also classified as IgM or IgA negative by each of the antigens included in the Luminex panel, with 8 exceptions that correspond to 3 samples (**Figure 1 and Table 2**). On the contrary, although a proportion of IgM/A positive samples classified by ELISA were also classified as positive by each of the antigens included in the Luminex panel, many of them were classified as IgM or IgA negative by Luminex (**Figure 1 and Table 2**).

The percentage of agreement for positive samples ranged from 4.79% for N CT to 16.44% for N FL, and from 8.90% for N CT to 21.23% for S, for IgM and IgA, respectively. The percentage of agreement for negative samples ranged from 15.19% for S2 to 17.01 % for N FL and from 15.60% for S to 17.12% for N FL, for IgM and IgA, respectively (**Table 2**). In this case, the antigens from the Luminex panel with the highest percentage of both positive and negative agreement were N FL for IgM and N FL and S2 for IgA (**Table 2**).

Considering the classification obtained by the combination of all the antigens included in the Luminex multiplex panel, the positive agreement was 23.29% for IgM and 30.14 % for IgA, while the negative agreement was 17.65% for IgM and 17.74% for IgA (**Table 2**). As a consequence, there were 113 (LM-E+: 109 mothers and 3 cords; LM+E-: 1 mother) and 105 (LM-E+: 99 mothers and 3 cords; LM+E-: 3 mothers) discrepant samples for IgM and IgA, respectively (**Figure 1** and **Table 2**). Among the LM-E+ discrepant samples from mothers, 95 out of 109 for IgM and 86 out of 99 for IgA were asymptomatic and either PCR negative or were not PCR tested. For both IgM and IgA, 1 sample was PCR positive and asymptomatic, and the remaining discrepant samples were all symptomatic with either positive (N=3 for IgM and N=3 for IgA), negative (N=8 for IgM and N=7 for IgA) or not done PCR (N=2 for IgM and N=2 for IgA). The only LM+E-sample for IgM was asymptomatic and PCR negative. Among the 3 LM+E-samples for IgA there were 2 asymptomatic, one with a negative PCR and one without PCR, and 1 symptomatic with a negative PCR. The 3 cord samples had discrepant results. There were no detectable levels of specific IgM and IgA against any of the antigens measured by Luminex, while ELISA gave a positive result. The corresponding mothers were: 1 LM+E+ for both IgM and IgA, 1 LM-E- for IgM and LM+E- for IgA, and 1 LM+E- for both IgM and IgA. In normal pregnancies, transplacental transfer of IgA is very limited and IgM does not cross the placenta (13), but in cases with severe infections significant levels of IgA and IgM can cross the placenta (15). In this study, none of the 3 mothers had severe COVID-19, which, together with the Luminex negative results, suggests that the results obtained by ELISA are false positive. However, vertical transmission of SARS-CoV-2 and the subsequent induction of immunoglobulins in the fetus should not be completely ruled out in cases of positive mothers, since transplacental transmission has been demonstrated in a severe COVID-19 pregnant case (16).

Regarding the performance of ELISA (**Table 3**) using Luminex results as gold standard, the SE was 97% for IgM and 94% for IgA, while the SP was as low as 18% for both IgM and IgA due to the many false positive results. As for the clinical performance parameters, PPV was 23% for IgM and 30% for IgA, while the NPP was 96% for IgM and 88% for IgA. Regarding the performance of ELISA based on subjects with a PCR test done, IgM/A reached a 100% SE while the SP was as low as 23% due to the many false positive. As for the PPV and NPP, the percentages were 15% and 100%, respectively.

The vast majority of discrepant samples (LM-E+) were asymptomatic and either PCR negative or not tested. In other words, the pretest probability of infection was very low and supports the classification of these discrepant samples as false positive results. At the moment, there is no recognized standard serological test to assess the sensitivity and specificity of other serological assays. Generally, neutralization assays and PCR test have been considered as the gold standard in other studies (17–23), but other assays such as CLIA have also been used (24). In this case, the use of PCR results as gold standard showed a very similar technical performance (SE and SP) compared to that obtained by using Luminex as stated above, although the set of samples used was slightly different since not all samples were from individuals with a PCR test done.

### Highly imbalanced technical and clinical performance for ELISA

Considering the results based on seropositivity by any of the isotypes included in the immunoassays (**Table 3**), the calculation of technical performance metrics resulted in a highly imbalanced performance for ELISA, with 96% SE at the expense of a 22% SP. As for the clinical performance, the NPV reached 87% while de PPV was 51%.

When prevalence of infection is low, the PPV of a test strongly relies on a high SP. For example, given a 95% SE and 5% prevalence, a decrease in SP from 100% to 95% would result in PPV dropping from 100% to 50%. On the contrary, in a scenario of 95% SE and 20% prevalence, a decrease in SP from 100% to 95% would result in a less steep fall of PPV from 100% to 83% (25). Conversely, changes in SE within the same context of prevalence do not affect so markedly the PPV and NPV.

In the case of ELISA, the low SP and PPV that we report here has important implications if this test is to be used in hospitals for their screenings. In particular, a 51% PPV implies that almost half of the people with a positive serological test would have not had passed the disease. Therefore, besides the overestimation of exposure to the virus, this would have an impact on the behavior of the people. Although people with positive serological tests are advised to keep the security measures, it is likely that they get a feeling of (false) protection and expose themselves more easily to the virus, increasing the chances of infection. In addition, false positive diagnosis may lead to unnecessary treatment and psychological distress. To avoid false positive results and their implications, if more sensitive and specific tests are not available, the pretest probability of infection should be taken into account when interpreting the results. In the case of serological tests, this includes symptomatology, contact with COVID-19 confirmed cases, previous history of COVID-19, presence of a PCR positive test and probability of alternative diagnosis (26).

Other reports have evaluated the performance of ELISA IgG and IgM/A from Vircell and have demonstrated diverse performances. Regarding IgG SE, different studies report the following ranges of: 65% (20), 86% (23), 13-93% (27), 36-93% (22), and 71-100% (21). The SE increased with days since onset of symptoms and disease severity (21,22,27). In our case, the SE for IgG does not reach the highest value reported by any of these papers.

A limitation of this study was the impossibility to stratify by days since onset of symptoms due to the small sample size. Ideally, performance validation should be done stratifying by days since onset of symptoms due to the kinetics of antibody production, which is delayed with respect to the onset of the infection. This is especially relevant in the case of IgG, which is the last immunoglobulin to develop during the course of an immune response. Therefore, the performance of a test highly depends on the time passed since the onset of infection, which can be monitored by the onset of symptoms or time since positivity of PCR in asymptomatic cases. In fact, our Luminex assay and others have demonstrated that performance reaches excellent levels at >10 o >14 days since onset of symptoms (6,21,22,27). Therefore, IgG SE assessment in this report would probably be higher had we stratified by days since onset of symptoms.

Concerning IgG SP, the same studies report the following ranges: 53% (22), 90% (23), 83-95% (21), 96% (20) and 97% (27). None of these values are as high as the 99% SP reported here, although most of them reach a considerably high SP.

With reference to IgM/A, the reported SE are as follows: 76-96% (22), 29-100% (27) and 77% (20). In this case, SE also increased with days since onset of symptoms and disease severity (22,27). The SE that we report here for IgM/A is concordant with the highest levels of those results. As for the SP, only one study reports a relatively high SP of 83% (20), while the other two inform a SP of 23% (22) and 46% (27). These values are not as low as what we report here but also show a very poor performance due to an elevated number of false positive samples. In fact, IgM is well-known for being a source of false positive results in immunoassays for many other infectious diseases due to its high non-specific reactivity, caused by cross-reactivity with other pathogens and the formation of rheumatoid factor (28). However, the latter is solved in the ELISA IgM/A from Vircell by the use of an IgG sorbent that removes rheumatoid factor complexes.

One explanation for the disagreement observed in some cases could be the source of biological samples. One study has reported an unsatisfactory performance of an immunochromatographic IgM/IgG rapid test in pregnant women due to an elevated false positive rate, and argue that the complexity of the immunological changes during pregnancy might be the underlying reason (24). Additionally, detection of cross-reactive antibodies generated by other coronaviruses or infectious diseases has been described and could be a source of false positive results not only for IgM (14,22).

In conclusion, we show a very low SP for the ELISA IgM/A compared to the high values reported by Vircell. In addition, the SE for the ELISA IgG was also lower than expected, although this value would probably be higher if only samples with >14 days since onset of symptoms were considered. Our results stress the need for highly specific and sensitive assays and external validation of diagnostic tests with different sets of samples.

## Data Availability

The authors confirm that the data supporting the findings of this study are available within the article. Raw data of this study are available from the corresponding author R.S. on request.

## Acknowledgements

We thank the Biobanks of Fundació Sant Joan de Déu, Clínic-IDIBAPS and Sant Pau for valuable management of samples, and the Centre for Genomic Regulation in Barcelona for the analysis of nasopharyngeal samples and the provision of the S protein. We are indebted to the nursing staff at Hospital Clínic and to the patients for their generous donation, to Fundació Glòria Soler for its support to the COVIDBANK initiative and to the HCB-IDIBAPS Biobank for the biological human samples and data procurement. We are grateful to F. Krammer for donation of RBD protein.

